# Occupational Risk Profiles for Infectious Disease Transmission in the Philippines: A Data-Driven Clustering Analysis

**DOI:** 10.64898/2026.05.07.26352625

**Authors:** Norvin P. Bansilan, Louie F. Dy, Jomar F. Rabajante

## Abstract

Occupational activities play a critical role in shaping patterns of infectious disease transmission, as work-related contact, exposure duration, and environmental conditions vary substantially across jobs. Evidence from infectious disease outbreaks, including tuberculosis and other respiratory infections, shows that occupations involving frequent close contact and crowded settings face elevated exposure risk, highlighting the need for systematic occupational risk assessment to inform public health action. This study adopts a data-driven approach to characterize occupational risk for infectious disease transmission in the Philippine workforce. Using job-specific indicators such as encounter frequency, work shift duration, and crowd density, unsupervised clustering methods were applied to group occupations into distinct risk profiles. Unlike predefined sector-based classifications, this approach identifies latent groupings that capture shared exposure characteristics and socioeconomic context. The resulting clusters reveal a clear gradient in exposure risk across occupations, with high-risk roles often concentrated among jobs with lower to moderate income levels, while lower-risk occupations tend to be associated with higher income and greater structural capacity for risk mitigation. These findings provide a framework for occupational risk stratification that is directly relevant for public health planning. Overall, this study demonstrates the value of integrating occupational and socioeconomic data to support targeted workplace interventions, risk-informed surveillance, and more equitable allocation of public health resources for infectious disease prevention and control in the Philippine context.

## 1. Introduction

Infectious disease transmission in workplace settings remains a significant public health concern because occupational activities shape patterns of human contact, exposure duration, and environmental risk, which are key determinants of disease spread. Work tasks influence how frequently individuals interact, the proximity of these interactions, and the environments in which they occur, thereby affecting transmission potential (Acke et al., 2022; Ghoroubi et al., 2023; Garus-Pakowska, 2022). Evidence shows that occupations involving sustained close contact, high encounter rates, or crowded conditions are associated with higher exposure risk compared to jobs with limited interpersonal interaction (Houštecká et al., 2021; Beale et al., 2022; Baharom et al., 2024). As such, occupational risk stratification plays an important role in informing disease prevention, enabling targeted interventions, risk-informed surveillance, and efficient allocation of public health resources.

In the Philippines, occupational risk assessment is shaped by a highly heterogeneous labor market, characterized by diverse work environments, varying levels of regulation, and significant socioeconomic disparities (PSA, 2021; PSA, 2025). Existing studies on occupational safety and health have primarily focused on workplace injuries, general occupational diseases, and policy implementation using descriptive or qualitative approaches (Faller et al., 2018; Lu, 2021; Conda et al., 2025). While these studies provide important insights, they largely emphasize observed outcomes rather than the exposure dynamics that drive infectious disease transmission. Furthermore, reliance on broad sector-based classifications may obscure substantial variation in risk across specific job roles. Consequently, there remains limited work that quantitatively characterizes infectious disease transmission risk across occupations using data-driven approaches that integrate both exposure and socioeconomic dimensions.

Recent advances in data availability and unsupervised machine learning methods offer an opportunity to address these limitations by enabling multivariate and data-driven occupational risk profiling (Hastie et al., 2009; Jain, 2010). In this study, we apply and compare multiple clustering algorithms, including K-means, hierarchical clustering, spectral clustering, Gaussian mixture models, and DBSCAN, to group Philippine occupations based on infectious disease spread risk and average monthly income. Cluster validity is assessed using established metrics, namely the Silhouette Score, Davies–Bouldin Index, and Calinski–Harabasz Index (Rousseeuw, 1987; Davies & Bouldin, 1979; Calinski & Harabasz, 1974). By identifying groups of occupations that share common exposure characteristics and socioeconomic vulnerability, this study provides a data-driven framework to support targeted workplace health interventions, risk-informed policy planning, and equitable allocation of public health resources in the Philippine context.

## 2. Methods

### 2.1 Data source and study variables

We compiled an occupational dataset from the *Jobs Risk Profiling: Philippines* interactive dashboard (Rabajante et al., n.d.), which integrates publicly available data from the Philippine Standard Industrial Classification (PSIC), Philippine Standard Occupational Classification (PSOC), O*NET Online, the Department of Labor and Employment, the Department of Budget and Management, and the Philippine Statistics Authority. The dataset includes 986 occupations and provides job-level information on infectious disease risk and average monthly income. Two variables were used in the clustering analysis: (1) the overall infectious disease spread risk score and (2) average monthly income, which serves as a proxy for socioeconomic vulnerability and capacity to adopt risk-mitigating measures. The risk score implemented in the dashboard is based on the infection risk modeling framework proposed by Dy and Rabajante (2020), which quantifies occupational exposure using contact-related and environmental factors.

### 2.2 Job Risk Calculator and risk score computation

The Job Risk Calculator operationalizes the Dy and Rabajante framework by incorporating four key determinants of infection risk: encounter rate, work shift duration, crowd density, and protection level. Encounter rate refers to the number of individuals a worker interacts with per hour, while work shift duration represents the total exposure time during a workday. Crowd density reflects how crowded the work environment is, and protection level captures the effectiveness of protective measures such as personal protective equipment, hygiene practices, and engineering controls. Together, these components represent the intensity, duration, and mitigation of workplace exposure.

### 2.3 Risk score formulation and point assignment

The overall occupational risk score is computed as:

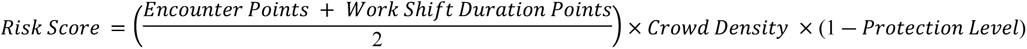

where encounter rate and work shift duration are first converted into standardized point values using predefined mappings. These point assignments enable heterogeneous occupational characteristics to be expressed on a comparable scale before integration with crowd density and protection level. This formulation reflects the combined effects of exposure frequency, duration, environmental conditions, and protective measures on infection risk, consistent with the modeling approach described by Dy and Rabajante (2020).

### 2.4 Unsupervised clustering analysis

All analyses were conducted using R within the RStudio integrated development environment (version 2024.09.0). We applied multiple unsupervised clustering methods to group occupations based on their combined profiles of infectious disease risk and monthly income. These approaches allow occupational groups with similar patterns of exposure and socioeconomic conditions to be identified directly from the data, without relying on predefined classifications.

To ensure that the resulting groupings were robust and not dependent on a single method, we implemented five clustering approaches representing different ways of identifying patterns in data, including partition-based, hierarchical, graph-based, model-based, and density-based techniques. The use of multiple methods allows for a more comprehensive assessment of occupational risk profiles and helps identify consistent groupings that are meaningful for public health interpretation.

#### 42.4.1 K-means clustering

K-means clustering groups occupations by identifying sets of jobs that are most similar to each other in terms of risk and income. Each occupation is assigned to a cluster such that occupations within the same group share comparable exposure and socioeconomic characteristics. This method is commonly used to identify relatively distinct and well-separated groups in population data.

#### 2.4.2 Hierarchical clustering

Hierarchical clustering organizes occupations into a tree-like structure based on their similarity. Starting from individual occupations, groups are gradually combined to form larger clusters. This approach is useful for examining how occupational risk profiles are related at different levels of similarity and for identifying broader groupings that may be relevant for policy and planning.

#### 2.4.3 Spectral clustering

Spectral clustering identifies occupational groups by examining relationships between occupations rather than relying solely on direct distances between variables. This approach is particularly useful when occupational risk profiles are not clearly separated and may follow more complex patterns, allowing for the detection of clusters that may not be captured by simpler methods.

#### 2.4.4 Gaussian mixture models

Gaussian mixture models (GMM) group occupations by estimating the probability that each occupation belongs to different clusters. This allows occupations to have partial membership across groups, which is useful in situations where risk profiles overlap. Such an approach reflects the reality that some occupations may share characteristics of multiple risk categories.

#### 2.4.5 DBSCAN

DBSCAN identifies clusters based on areas where occupations are densely concentrated in terms of similar risk and income profiles. It also distinguishes occupations that do not belong to any clear group, which may represent unique or outlier risk conditions. This is useful for identifying both common occupational profiles and atypical cases that may require special attention.

### 2.5 Cluster validation and evaluation

Because clustering does not rely on predefined categories, we evaluated the quality of the resulting occupational groupings using internal validation measures that assess how well occupations are grouped and how distinct the groups are from one another. These measures were applied across all clustering methods to assess the consistency and robustness of the results.

#### 2.5.1 Silhouette Score

The Silhouette Score measures how closely each occupation matches its assigned group relative to other groups. Higher values indicate that occupations are well matched to their cluster and clearly separated from others, suggesting more meaningful groupings.

#### 2.5.2 Davies–Bouldin Index

The Davies–Bouldin (DB) Index assesses how similar each group is to other groups based on the spread of occupations within clusters and the separation between clusters. Lower values indicate better-defined and more distinct occupational groupings.

#### 2.5.3 Calinski–Harabasz Index

The Calinski–Harabasz (CH) Index evaluates how well clusters are separated relative to how tightly occupations are grouped within each cluster. Higher values indicate stronger clustering structure and more clearly defined occupational profiles.

## 3 Results

### 3.1 Cluster validation and evaluation

Figure 1 presents the internal validation metrics for each clustering method (K-means, hierarchical, spectral, and GMM) across different values of *k* and epsilon (*ε*) for DBSCAN, including the Silhouette Score, DB Index, and CH Index. These metrics evaluate clustering quality by how well occupations are grouped and how distinct the clusters are from one another. A summary table of all the indices for each clustering method is shown in the Appendix Table A1.

**Figure 1.**
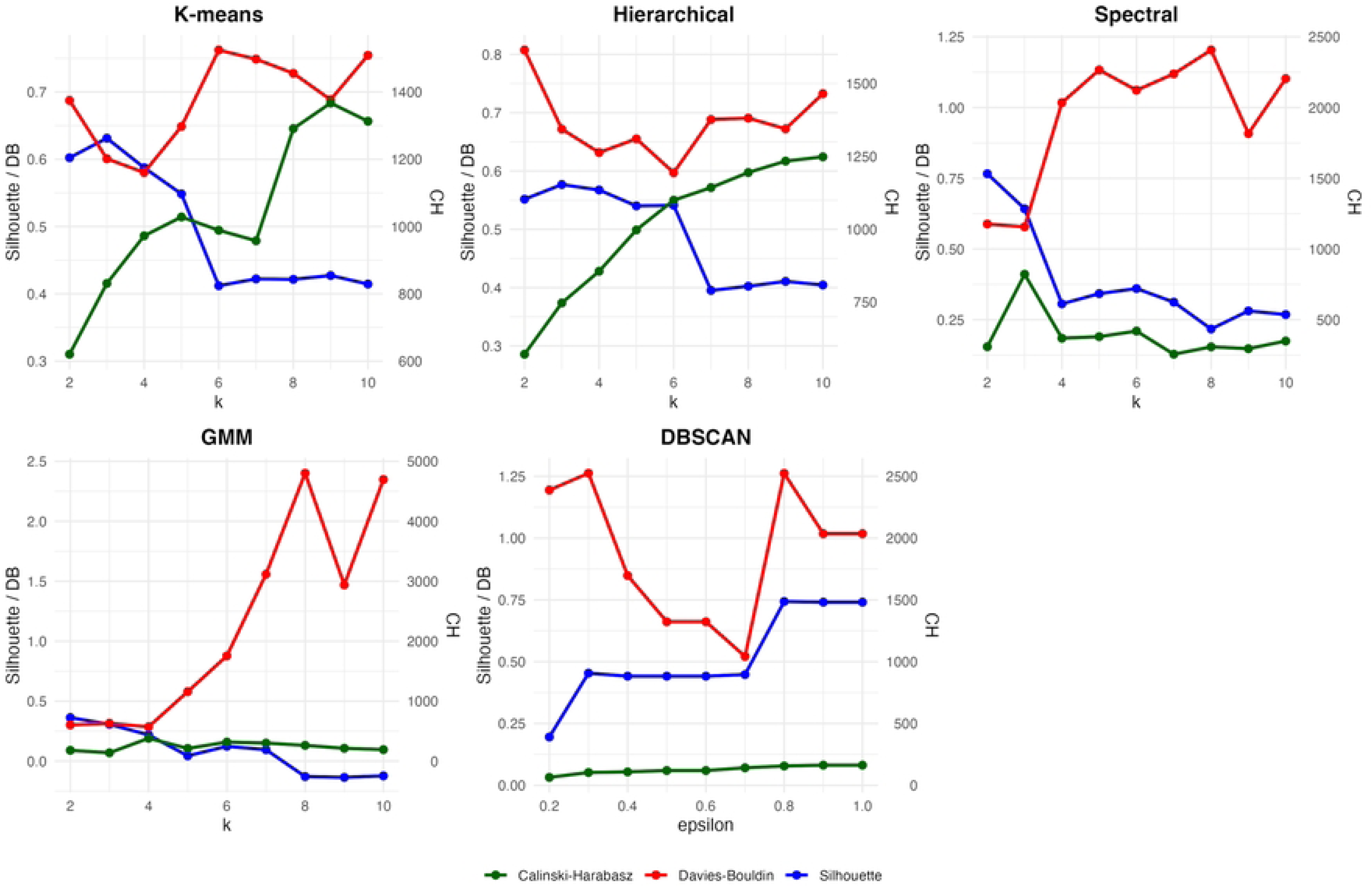
Panels show results for each clustering method across varying numbers of *k* or *∈*. Three metrics are presented: CH Index (green), DB Index (red), and Silhouette Score (blue). The left vertical axis corresponds to the Silhouette Score and DB Index, while the right vertical axis corresponds to the CH Index.

For K-means clustering, *k* = 4 was selected as the optimal number of clusters. At lower values (*k* < 4), clusters were insufficiently differentiated, indicating limited ability to distinguish occupational risk profiles. At higher values (*k* > 4), although clusters appeared more distinct, the separation between clusters decreased, resulting in more homogeneous groupings overall.

For hierarchical clustering, *k* = 6 was identified as the most appropriate solution. When fewer clusters were specified (*k* < 6), the resulting groups lacked sufficient differentiation. Increasing the number of clusters beyond this point (*k* > 6) led to reduced separation, suggesting over-fragmentation of occupational profiles without meaningful gains in interpretability.

For spectral clustering, *k* = 3 was determined to be the most suitable configuration. At lower values, clusters were separated but not well differentiated, while higher values resulted in clusters that were neither clearly distinct nor well separated. In addition, consistently low Silhouette and CH values, together with relatively high DB values, indicate weak clustering structure, suggesting that spectral clustering is not well-suited for this dataset.

For GMM, *k* = 4 was selected as the optimal number of components. However, minimal variation in validation metrics was observed for *k* < 4, indicating limited sensitivity to cluster number. At higher values (*k* > 4), clustering performance deteriorated, with reduced separation and coherence. Similar to spectral clustering, the generally low Silhouette and CH values and higher DB values suggest that GMM provides less well-defined occupational groupings.

For DBSCAN, the most meaningful clustering structure was observed at *ε* = 0.7, which produced three clusters. As the value of *ε* increased, the number of identified clusters decreased, indicating that occupations were increasingly grouped into broader, less distinct categories. Specifically, smaller values of *ε* (e.g., 0.2-0.3) resulted in a larger number of clusters (5-7 clusters), reflecting fragmented groupings with limited cohesion. Moderate values (*∈* = 0.4 ― 0.6) produced four clusters, suggesting improved consolidation but still limited separation. At higher values (*ε* ≥ 0.8), the number of clusters decreased to two, indicating overgeneralization of occupational profiles.

Overall, K-means and hierarchical clustering demonstrated more stable and interpretable clustering structures compared to the other methods and were therefore prioritized for subsequent analysis.

### 3.2 Clustering methods

The application of K-means clustering (*k* = 4) resulted in distinct groupings of occupations based on their combined profiles of infectious disease exposure risk and average monthly income, see Figure 2. The characteristics of each cluster are described below.

**Figure 2.**
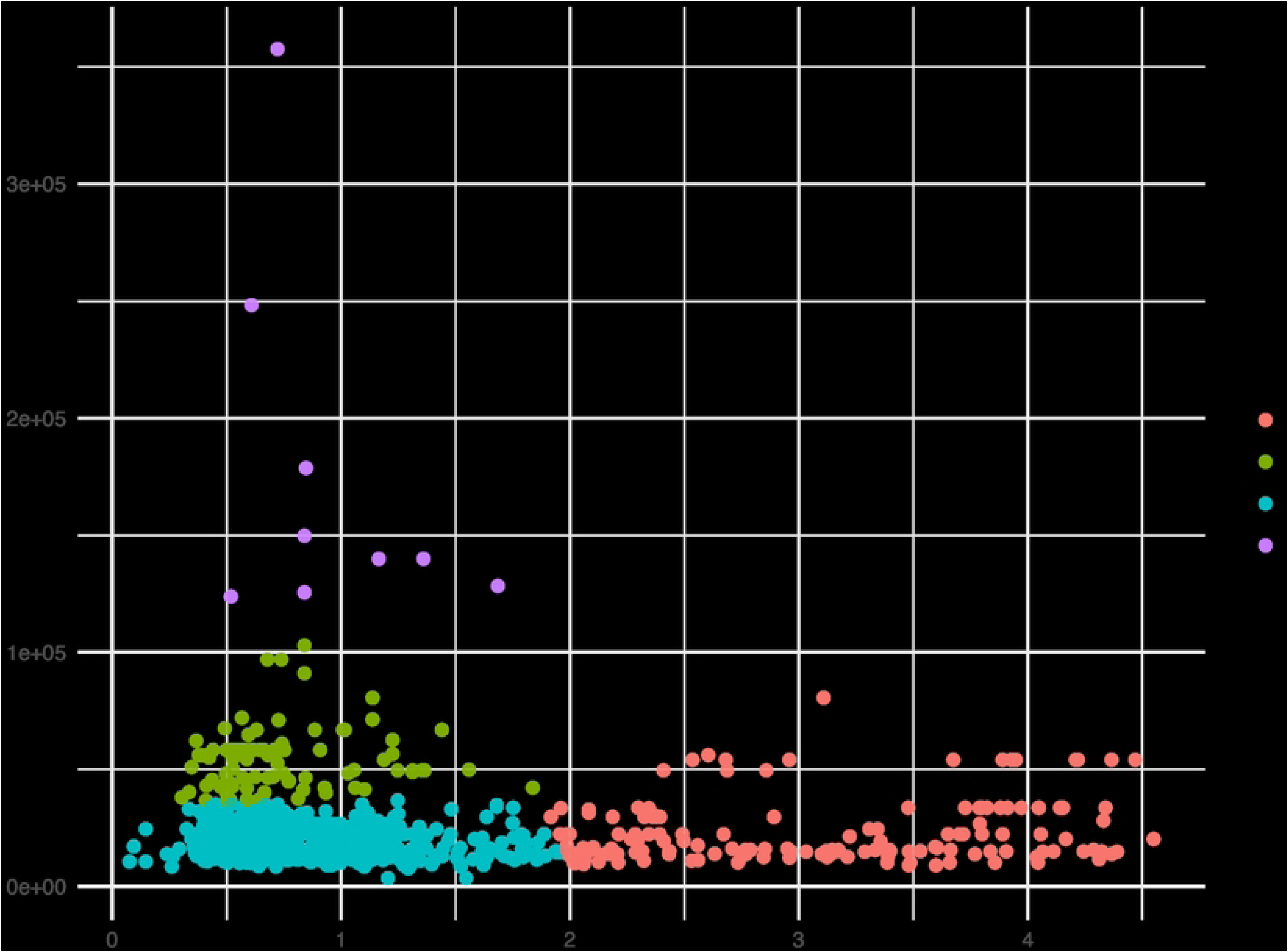
K-means clustering results (*k* = 4) for Philippine occupations based on overall infectious disease risk score and average monthly income. Each point represents an occupation, colored according to cluster membership. The horizontal axis shows the overall risk score, reflecting exposure potential, while the vertical axis represents average monthly income.

Cluster 1 (red) represents occupations characterized by high infectious disease exposure risk, as indicated by elevated risk scores, combined with generally low to moderate income levels. This cluster is predominantly composed of healthcare and clinical occupations, including physicians, nurses, and allied health professionals, whose roles involve direct and prolonged patient contact. In addition, the cluster includes emergency response and public safety personnel, such as firefighters, police officers, and emergency medical workers, who operate in high-contact and often unpredictable environments. A substantial portion of the cluster also consists of caregiving and personal support roles, including home health aides and childcare workers, which require sustained close interaction with individuals. Furthermore, health-related technical occupations, such as laboratory technicians and imaging specialists, are represented, reflecting exposure through handling biological materials and clinical procedures. The cluster also captures high-contact service and regulatory roles, including teachers, transport workers, and inspectors, where frequent interaction with large or dynamic populations increases transmission potential.

Cluster 2 (green) represents occupations characterized by moderate infectious disease exposure risk combined with relatively higher income levels, reflecting roles that are more knowledge-intensive and less reliant on sustained close interpersonal contact. This cluster is predominantly composed of professional, technical, and managerial occupations, including engineers (e.g., civil, electrical, mechanical, and environmental engineers), scientists (e.g., economists, statisticians, chemists), and information technology specialists such as software developers and computer systems analysts. A substantial portion of this group also includes managerial and administrative roles, such as financial managers, human resource managers, supply chain managers, and construction managers, which typically involve structured work environments with controlled interactions. In addition, regulatory, planning, and oversight occupations, including urban planners, compliance managers, and policy-related roles, are represented, reflecting responsibilities that involve coordination rather than direct service delivery. Some specialized operational and supervisory roles, such as air traffic controllers and engineering managers, are also included, where interaction is present but often mediated through systems or protocols.

Cluster 3 (blue) represents occupations characterized by moderate to high variability in infectious disease exposure risk and generally low to moderate income levels, reflecting a diverse mix of job roles that combine manual labor, service-oriented tasks, and technical functions. This cluster includes a large proportion of skilled trades and industrial occupations, such as construction workers, machine operators, mechanics, and equipment technicians, where exposure risk arises from shared workspaces, physical proximity, and variable adherence to protective measures. It also encompasses transportation, logistics, and operational roles, including drivers, dispatchers, and warehouse-related occupations, which involve frequent interaction with goods, environments, and, in some cases, the public. In addition, a significant number of service and customer-facing occupations are represented, such as food service workers, retail staff, hospitality personnel, and personal care providers, where sustained interpersonal contact increases transmission potential. The cluster further includes administrative, clerical, and support roles, as well as creative, educational, and technical occupations, reflecting heterogeneity in both exposure patterns and workplace environments.

Cluster 4 (purple) represents occupations characterized by low infectious disease exposure risk and high income levels, reflecting roles that are typically performed in controlled, structured, or highly managed environments with limited routine close contact. This cluster is composed primarily of senior leadership, executive, and high-level administrative positions, including chief executives, general managers, and high-ranking military officials, whose responsibilities are largely strategic and supervisory rather than operational. It also includes judicial and decision-making roles, such as judges and administrative law officers, where interactions are formalized and often occur in regulated settings. Additionally, specialized high-skill occupations, such as airline pilots, are present, where interaction with others is limited or mediated through protocols and controlled environments. The inclusion of executive administrative roles further reflects occupations that involve coordination and communication, but typically within structured and less crowded settings.

On the other hand, the application of hierarchical clustering (*k* = 6) provides a more granular segmentation of occupations based on their combined profiles of infectious disease exposure risk and average monthly income, see Figure 3. Compared to the partition-based K-means approach, hierarchical clustering captures nested and more nuanced relationships among occupations, allowing for finer distinctions in both exposure patterns and socioeconomic positioning. The resulting clusters highlight additional heterogeneity within occupational groups, particularly among moderate-risk and lower-income categories. The characteristics of each cluster are described below.

**Figure 3.**
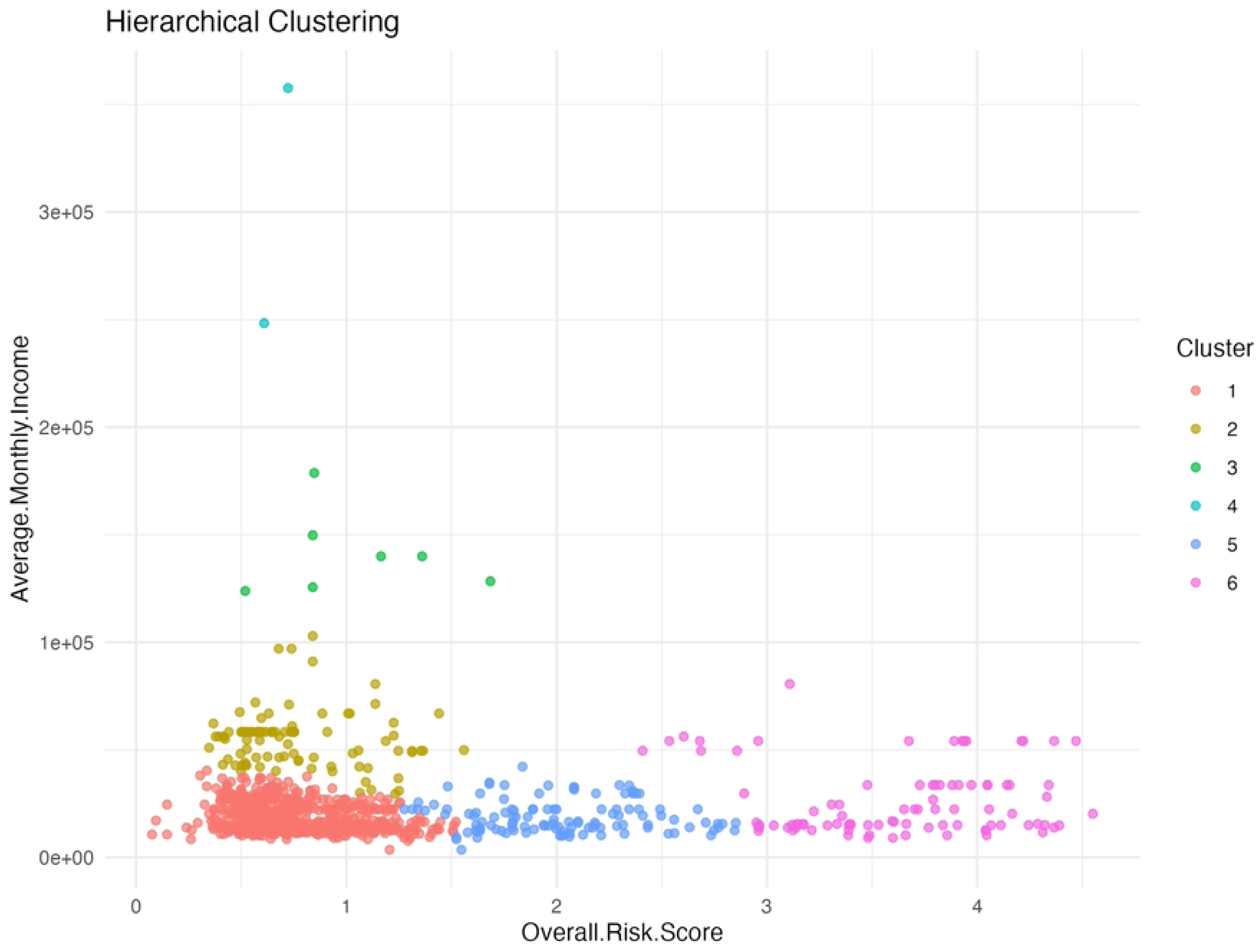
Hierarchical clustering results (*k* = 6) for Philippine occupations based on overall infectious disease risk score and average monthly income. Each point represents an occupation, colored according to cluster membership derived from an agglomerative clustering approach using Ward’s linkage. The horizontal axis represents the overall risk score, while the vertical axis shows average monthly income.

Cluster 1 (red) represents a large and heterogeneous group of occupations characterized by generally low to moderate infectious disease exposure risk and predominantly low to moderate income levels. This cluster encompasses a wide range of manual labor, technical, service, and creative occupations, reflecting variability in both workplace environments and exposure conditions. A substantial portion of this group consists of skilled trades and industrial workers, including machine operators, construction laborers, mechanics, and manufacturing-related occupations, where exposure risk is influenced by shared workspaces and physical proximity but often lacks sustained close interpersonal interaction. The cluster also includes service and customer-facing roles, such as retail workers, food service staff, and hospitality personnel, where intermittent public interaction contributes to moderate exposure risk. In addition, a notable presence of technical, scientific, and analytical occupations is observed, including engineers, data specialists, environmental scientists, and IT-related roles, which are typically performed in more controlled environments with lower direct contact intensity. Creative and educational occupations, such as artists, writers, and teachers, are also represented, further contributing to the heterogeneity of the cluster (Appendix Figure A6).

Cluster 2 (yellow) is composed predominantly of professional, technical, and managerial occupations characterized by moderate infectious disease exposure risk and relatively higher income levels compared to other clusters. This group is largely composed of engineers, analysts, scientists, and corporate or administrative managers, including roles in energy systems, information technology, finance, logistics, and infrastructure planning. Many of these occupations are performed in structured and controlled work environments, such as offices, laboratories, or specialized industrial settings, where exposure to infectious disease is generally limited by reduced frequency of close interpersonal contact and the presence of organizational safety protocols. In addition, the cluster includes high-level administrative and leadership roles, such as managers, directors, and military officers, as well as specialized professionals in healthcare and research, including pathologists and clinical research coordinators (Appendix Figure A7).

Cluster 3 (green) represents a small but distinct group of high-ranking, executive, and decision-making occupations characterized by low infectious disease exposure risk and high income levels. This cluster includes senior leadership and governance roles, such as generals, high-level military officers, chief executives, and general and operations managers, as well as judicial and adjudication positions, including judges and administrative law officials. These occupations are typically performed in highly controlled, formalized environments with limited routine, close interpersonal contact, where interactions are structured, regulated, and often mediated through administrative processes. The presence of airline pilots in this cluster further reflects specialized roles with controlled operational settings, where exposure is mitigated through protocols, restricted access environments, and institutional safeguards (Appendix Figure A8).

Cluster 4 (cyan) represents a very small and highly specialized group of occupations characterized by low infectious disease exposure risk and high income levels, consisting primarily of executive and high-level administrative roles such as chief executives and assessors. These occupations are typically performed in controlled, office-based or institutional environments, where interactions are structured, limited in frequency, and often involve decision-making rather than direct service or operational tasks. As a result, opportunities for sustained close contact are relatively minimal, contributing to lower exposure risk (Appendix Figure A9).

Cluster 5 (blue) represents a large and diverse group of occupations characterized by moderate to high infectious disease exposure risk and generally low to moderate income levels, reflecting roles that involve frequent interpersonal interaction and service-oriented responsibilities. This cluster includes a substantial number of frontline service and care-related occupations, such as healthcare support workers, counselors, social workers, childcare providers, and personal care aides, where exposure risk is elevated due to sustained close contact and direct interaction with individuals in community or institutional settings. It also encompasses public-facing and customer service roles, including cashiers, food service workers, transportation operators, and retail-related occupations, where repeated interactions with the public contribute to increased transmission potential. In addition, the cluster includes skilled trades, maintenance, and operational roles, such as electricians, repair workers, plumbers, and technicians, where exposure may arise from shared workspaces and variable adherence to protective measures. A notable presence of protective service and regulatory occupations, including police personnel, investigators, and correctional staff, further reflects roles involving direct engagement with the public in dynamic and often uncontrolled environments. Educational and allied health professions are also represented, contributing to the diversity of exposure settings within the cluster (Appendix Figure A10).

Cluster 6 (purple) represents occupations characterized by high infectious disease exposure risk and a wide range of income levels, with a strong concentration of healthcare, emergency response, and close-contact service professions. This cluster is dominated by clinical and allied health occupations, including physicians, nurses, specialists, therapists, and diagnostic technologists, where exposure risk is inherently elevated due to direct and sustained contact with patients, biological samples, and healthcare environments. It also includes emergency and protective service roles, such as firefighters, paramedics, correctional officers, and law enforcement personnel, where interactions occur in dynamic and often unpredictable settings with limited control over exposure conditions. In addition, the cluster contains personal care and close-contact service occupations, such as barbers, dental assistants, and therapy aides, where proximity to individuals further increases transmission potential. The presence of aviation-related roles, such as flight attendants, highlights occupations involving high mobility and repeated exposure to diverse populations, further contributing to risk. Unlike other clusters, this group spans a broader income spectrum, ranging from support-level healthcare workers to highly specialized medical professionals, indicating that high exposure risk is not confined to lower-income occupations but extends across different levels of professional hierarchy (Appendix Figure A11).

Additional clustering approaches, including GMM, spectral clustering, and DBSCAN, were also explored to examine alternative structures in the data. The results from these methods are presented in the Appendix Figures A4, A5, and A6.

## 4 Discussion

This study demonstrates the value of unsupervised machine learning in identifying distinct occupational risk profiles for infectious disease transmission in the Philippine context. By integrating exposure-related indicators with socioeconomic information, the analysis shows that occupational risk is not uniformly distributed across the workforce, but instead reflects systematic differences in job function, work environment, and economic capacity.

Across the clustering methods evaluated, K-means and hierarchical clustering consistently produced more stable and interpretable solutions, as supported by internal validation metrics. These methods achieved relatively higher Silhouette scores and CH indices, alongside lower DB values, indicating better-defined and well-separated clusters. In contrast, spectral clustering and GMM exhibited lower cluster cohesion and separation, while DBSCAN showed sensitivity to parameter selection, leading to unstable cluster structures and inconsistent cluster counts. These findings suggest that the occupational dataset is better represented by globally partitioned structures rather than density-based or probabilistic clustering assumptions, supporting the use of K-means and hierarchical clustering for policy-relevant interpretation.

Using these more robust methods, a clear gradient between exposure risk and income emerges across clusters. Results from K-means clustering reveal four distinct occupational groupings. Across the four clusters, a clear gradient emerges linking infectious disease exposure risk with socioeconomic positioning, revealing important structural patterns in the occupational landscape. Cluster 1 captures frontline and service-oriented occupations with high exposure risk and generally low to moderate income, highlighting groups that are both epidemiologically vulnerable and economically constrained. In contrast, Cluster 2 consists of professional, technical, and managerial roles characterized by moderate exposure risk and higher income, suggesting more controlled work environments and greater access to protective resources. Cluster 3 represents a heterogeneous group of occupations with variable exposure conditions and relatively lower income levels, reflecting workers in mixed environments where risk is influenced by both workplace setting and task variability. Finally, Cluster 4 comprises executive, leadership, and highly specialized roles with low exposure risk and high income, indicating strong capacity to mitigate risk through controlled environments and flexible work arrangements.

Taken together, these clusters illustrate that occupational exposure to infectious diseases is not evenly distributed across the workforce, but instead aligns with differences in job function, work environment, and economic capacity. This pattern underscores the need for targeted, occupation-specific public health interventions, as well as policies that address both exposure risk and structural vulnerability. In particular, the concentration of high-risk, lower-income occupations in Cluster 1 highlights a critical area for prioritizing workplace protections, surveillance, and resource allocation in the Philippine context.

In hierarchical clustering, there are six clusters formed. Cluster 1 reflects a broad segment of the workforce with mixed exposure profiles but relatively limited economic capacity, suggesting that while individual occupations may not consistently experience high exposure risk, structural factors such as workplace conditions and resource constraints may still influence vulnerability. The diversity within this cluster highlights the importance of context-specific risk assessment, as exposure potential varies substantially even among occupations grouped within similar income ranges. This cluster underscores that a significant portion of the workforce operates in conditions where exposure risk is neither negligible nor extreme, but shaped by variability in tasks, environments, and interaction patterns.

Despite the relatively lower direct exposure compared to high-contact occupations, cluster 2 reflects strategically important roles in economic, technological, and institutional systems, where workforce disruptions may have broader systemic implications. The combination of moderate exposure risk and higher socioeconomic status suggests a greater capacity for risk mitigation through remote work, access to protective measures, and organizational support. However, the diversity of roles within this cluster also indicates that exposure risk may vary depending on specific work settings, particularly for occupations involving fieldwork, supervision, or operational oversight. Overall, Cluster 2 highlights a group of occupations where infection risk is shaped more by environmental and organizational context than by sustained interpersonal contact, underscoring the need for differentiated workplace policies even within relatively advantaged sectors.

Cluster 3 highlights a group of occupations with both low exposure risk and substantial economic and organizational resources, indicating a strong capacity to mitigate infection risk through workplace control, access to protective measures, and flexible work arrangements. Compared to other clusters, these roles are less constrained by direct service or frontline interaction, reinforcing a broader pattern in which occupational hierarchy and socioeconomic status are associated with reduced vulnerability to infectious disease exposure.

Despite its small size, cluster 4 reflects occupations with significant economic and organizational influence, as well as strong access to resources that support risk mitigation, including flexible work arrangements, administrative control over work environments, and the ability to implement protective measures. The presence of these roles further reinforces the broader pattern observed across clusters, in which higher socioeconomic position is associated with reduced occupational exposure to infectious disease, highlighting structural inequalities in workplace risk distribution.

Cluster 5 captures a segment of the workforce that is highly interactive, service-driven, and operationally essential, yet often constrained by moderate income levels and limited control over workplace conditions. This combination of elevated exposure risk and constrained socioeconomic capacity suggests increased vulnerability, highlighting the importance of prioritizing these occupations in workplace safety policies, surveillance systems, and targeted public health interventions.

Cluster 6 captures the core high-exposure segment of the workforce, where infection risk is driven by the nature of work itself rather than workplace variability. These findings underscore the critical importance of prioritizing these occupations in infection prevention strategies, including access to protective equipment, vaccination programs, surveillance systems, and institutional safeguards, as well as ensuring sustained support for both frontline and specialized healthcare workers in managing infectious disease risks.

Across the six clusters identified through hierarchical clustering, a more nuanced and stratified pattern of occupational risk emerges, further reinforcing the interplay between infectious disease exposure, job function, and socioeconomic positioning. The results reveal a continuum of risk, ranging from low-exposure, high-income leadership roles in Clusters 3 and 4, to high-exposure occupations concentrated in healthcare, emergency response, and close-contact services in Cluster 6. Notably, Cluster 5 highlights a substantial segment of the workforce engaged in frontline service, care, and operational roles, where exposure risk is elevated, but economic capacity remains constrained. In contrast, Cluster 2 captures professional and technical occupations with moderate exposure risk and relatively higher income, reflecting structured environments with greater access to protective resources. Cluster 1, meanwhile, represents a broad and heterogeneous group with mixed exposure conditions, emphasizing that risk is not uniform even among occupations with similar income levels.

Taken together, these findings demonstrate that occupational exposure to infectious disease is shaped by both the nature of work and structural inequalities, with certain groups facing compounded vulnerability due to high exposure and limited economic resources. The additional granularity provided by hierarchical clustering highlights important within-group differences that are not as evident in partition-based approaches, particularly among moderate-risk occupations. This underscores the value of using more flexible clustering methods to uncover latent occupational risk profiles, which can inform more precise and equitable public health interventions. In the Philippine context, these results support the need for differentiated, occupation-specific strategies, including targeted protection for high-exposure groups, strengthened workplace safety policies for service-oriented sectors, and broader structural support for workers with limited capacity to mitigate risk.

In both approaches, occupations involving frequent interpersonal contact and prolonged exposure, such as healthcare workers, emergency responders, and high-contact service roles, consistently fall within high-risk clusters that are associated with lower to moderate income levels. These occupations are essential to public health and service delivery, yet often operate under conditions with limited control over exposure, highlighting a critical vulnerability.

At the opposite end of the spectrum, clusters composed of executive, leadership, and highly specialized professional roles are characterized by low exposure risk and high income. These occupations typically function in controlled or flexible environments, where interaction is limited or mediated through organizational structures. This pattern suggests that higher-income occupations are not only less exposed but also better positioned to implement and sustain protective measures, reinforcing structural inequalities in occupational health outcomes.

Intermediate clusters further reveal important nuances. Professional, technical, and managerial occupations tend to exhibit moderate exposure risk with relatively higher income, reflecting structured environments and access to institutional safeguards. Meanwhile, clusters composed of manual labor, service, and logistics-related roles show mixed exposure profiles but generally lower income levels, indicating that risk is influenced not only by occupation type but also by workplace conditions and task variability. The additional granularity observed in hierarchical clustering highlights that even within similar income groups, exposure risk can vary substantially, underscoring the importance of fine-grained occupational analysis.

These findings have important implications for public health policy. First, they highlight the limitations of broad sector-based classifications, which may mask substantial heterogeneity in exposure risk across occupations. The clustering framework developed in this study provides a more nuanced basis for targeted interventions, including prioritization of high-risk occupations for vaccination, personal protective equipment, surveillance, and workplace regulation. Second, the observed alignment between high exposure risk and lower income in several clusters underscores the need for equity-oriented policies, such as hazard compensation, strengthened occupational safety standards, and expanded social protection mechanisms.

From a methodological perspective, the use of multiple clustering approaches and validation metrics strengthens the robustness of the findings. The convergence of results across K-means and hierarchical clustering, combined with consistent metric performance, supports the reliability of the identified occupational groupings. At the same time, the weaker performance of spectral clustering, GMM, and DBSCAN highlights the importance of method selection in applied clustering studies, particularly when results are intended to inform policy decisions.

Several limitations should be considered. The risk scores used in this study are derived from a modeling framework based on generalized workplace characteristics and may not fully capture context-specific variations in behavior, compliance, or environmental conditions. In addition, the analysis relies on cross-sectional occupational data and does not account for temporal changes in workplace practices or disease dynamics. Future research could incorporate longitudinal data, real-world infection outcomes, and more detailed workplace indicators to further refine risk estimation and clustering accuracy.

Despite these limitations, this study provides a data-driven framework for occupational risk stratification that is directly relevant to public health planning in the Philippines. By identifying clusters of occupations with shared exposure and socioeconomic characteristics, the findings support more efficient and equitable allocation of resources and interventions. More broadly, this work demonstrates how machine learning methods can be effectively integrated with public health principles to address complex, multidimensional challenges in occupational health.

## 5 Conclusion

This study demonstrates that the occupational risk of infectious disease transmission in the Philippines is structured, unequal, and closely linked to both job characteristics and socioeconomic position. Using unsupervised machine learning, distinct clusters of occupations were identified, revealing a clear gradient in which high-exposure roles are often associated with lower income and limited capacity for risk mitigation, while higher-income occupations tend to experience lower exposure and greater structural protection.

The consistent performance of K-means and hierarchical clustering, supported by validation metrics, highlights their suitability for identifying policy-relevant occupational groupings. These findings underscore the importance of moving beyond broad sector-based classifications toward data-driven, occupation-level risk stratification.

From a public health perspective, the results provide a practical framework for targeted interventions, including prioritization of high-risk occupations for protective measures, improved workplace safety policies, and more equitable allocation of resources. Ultimately, this study illustrates how integrating machine learning with public health principles can support more precise, equitable, and evidence-based responses to infectious disease risk in the workforce.

## Data Availability

All relevant data are within the manuscript and its Supporting Information files.

https://drive.google.com/file/d/1-7MbcH1XuBdLntaIRJ7sp0l7Q-st7Kin/view?usp=sharing

